# The Value of Normal Interictal EEGs in Epilepsy Diagnosis and Treatment Planning: A Retrospective Cohort Study using Population-level Spectral Power and Connectivity Patterns

**DOI:** 10.1101/2025.01.03.25319963

**Authors:** Neeraj Wagh, Andrea Duque-Lopez, Boney Joseph, Brent Berry, Lara Jehi, Daniel Crepeau, Leland Barnard, Venkatsampath Gogineni, Benjamin H. Brinkmann, David T. Jones, Gregory Worrell, Yogatheesan Varatharajah

## Abstract

**Introduction:** Scalp electroencephalography (EEG) is a cornerstone in the diagnosis and treatment of epilepsy, but routine EEG is often interpreted as normal without identification of epileptiform activity during expert visual review. The absence of interictal epileptiform activity on routine scalp EEGs can cause delays in receiving clinical treatment. These delays can be particularly problematic in the diagnosis and treatment of people with drug-resistant epilepsy (DRE) and those without structural abnormalities on MRI (i.e., MRI negative). Thus, there is a clinical need for alternative quantitative approaches that can inform diagnostic and treatment decisions when visual EEG review is inconclusive. In this study, we leverage a large population-level routine EEG database of people with and without focal epilepsy to investigate whether normal interictal EEG segments contain subtle deviations that could support the diagnosis of focal epilepsy.

**Data & Methods:** We identified multiple epochs representing eyes-closed wakefulness from 19-channel routine EEGs of a large and diverse neurological patient population (N=13,652 recordings, 12,134 unique patients). We then extracted the average spectral power and phase-lag-index-based connectivity within 1-45Hz of each EEG recording using these identified epochs. We decomposed the power spectral density and phase-based connectivity information of all the visually reviewed normal EEGs (N=6,242) using unsupervised tensor decompositions to extract dominant patterns of spectral power and scalp connectivity. We also identified an independent set of routine EEGs of a cohort of patients with focal epilepsy (N= 121) with various diagnostic classifications, including focal epilepsy origin (temporal, frontal), MRI (lesional, non-lesional), and response to anti-seizure medications (responsive vs. drug-resistant epilepsy). We analyzed visually normal interictal epochs from the EEGs using the power-spectral and phase-based connectivity patterns identified above and evaluated their potential in clinically relevant binary classifications.

**Results:** We obtained six patterns with distinct interpretable spatio-spectral signatures corresponding to putative aperiodic, oscillatory, and artifactual activity recorded on the EEG. The loadings for these patterns showed associations with patient age and expert-assigned grades of EEG abnormality. Further analysis using a physiologically relevant subset of these loadings differentiated patients with focal epilepsy from controls without history of focal epilepsy (mean AUC 0.78) but were unable to differentiate between frontal or temporal lobe epilepsy. In temporal lobe epilepsy, loadings of the power spectral patterns best differentiated drug-resistant epilepsy from drug-responsive epilepsy (mean AUC 0.73), as well as lesional epilepsy from non-lesional epilepsy (mean AUC 0.67), albeit with high variability across patients.

**Significance:** Our findings from a large population sample of EEGs suggest that normal interictal EEGs of patients with epilepsy contain subtle differences of predictive value that may improve the overall diagnostic yield of routine and prolonged EEGs. The presented approach for analyzing normal EEGs has the capacity to differentiate several diagnostic classifications of epilepsy, and can quantitatively characterize EEG activity in a scalable, expert-interpretable, and patient-specific fashion. Further technical development and clinical validation may yield normal EEG-derived computational biomarkers that could augment epilepsy diagnosis and assist clinical decision-making in the future.

## 1. Introduction

Epilepsy is a neurological disorder characterized by recurrent, unprovoked seizures and is estimated to affect ∼50 million people worldwide^1^. A scalp electroencephalogram (EEG) non-invasively records the electrical activity of the brain, and its findings play a critical role in the clinical diagnosis and management of epilepsy^2–4^. The diagnostic yield of a short 20–40-minute routine EEG is determined by the presence of spontaneous transient interictal epileptiform discharges (IEDs)^5–7^. However, ∼30-55% of routine EEGs of patients with epilepsy and 9-10% of prolonged video EEGs show no evidence of IEDs and delay the diagnosis of epilepsy^12–17^.

In newly diagnosed epilepsy, anti-seizure medications (ASMs) are the first choice of therapy. However, despite a successful diagnosis, about half the patients do not respond to their first ASM, and about a third continue to have uncontrolled seizures despite multiple ASM trials^14,15^. Therefore, the determination of drug-resistant epilepsy (DRE) can take several months or years, while the patients continue to experience seizures and comorbidities. Thus, the early identification of DRE is essential to reduce disease burden and to initiate evaluations for additional therapies such as resective surgery and electrical brain stimulation. In focal epilepsy, magnetic resonance imaging (MRI) scans of the brain can help clarify the disease etiology by identifying structural abnormalities that lead to seizures^16^. In MRI negative, i.e., non-lesional, epilepsy patients, normal EEGs can cause further delays in identifying the epileptogenic brain regions for treatment. Broadly, the inability to identify interictal epileptiform activity during visual review of routine EEGs can delay the initiation of ASMs, increase healthcare costs^18^, and put the patient at an increased risk of seizure-related injuries and comorbidities^18,19^.

As such, there is a clear need for alternative approaches that can assist with early diagnosis and treatment planning when traditional routine EEG tests are inconclusive. Our goal in this study is to develop a quantitative approach to explore automatic analysis of normal interictal EEGs, which could provide early, objective, and inexpensive clinical decision support. Emerging evidence suggests that quantitative approaches based on expert EEG features and black-box machine learning models have the potential to improve the diagnostic value of routine EEGs and augment decision-making in epilepsy^17–24^. However, expert-defined features may not sufficiently capture the complexity of multivariate EEG activity and black-box models face significant robustness and interpretability issues. Building on prior work, here we take a data-driven and interpretable approach -- leveraging a large population database using unsupervised tensor decompositions -- to identify spectral power and connectivity patterns of normal interictal EEG and evaluate their potential in differentiating various focal epilepsy classifications.

In this study, we retrospectively analyzed a large dataset of 13,652 routine EEGs from a diverse neurological population of 12,134 adults and a cohort of 121 adults with confirmed focal epilepsy. Patterns of power spectral density and phase-based connectivity in eyes-closed wakefulness were extracted from the 6,242 normal EEGs in the population dataset using canonical polyadic tensor decomposition. We examined the spatial and frequency distributions of these patterns and investigated their association with age and clinically assigned EEG grades. Then, pattern loadings were computed to quantitatively characterize the normal EEG activity (i.e., interictal non-epileptiform) of patients with focal epilepsy. With these loadings, we studied group differences and conducted classification analyses to explore the use of normal EEGs in epilepsy diagnosis and treatment planning.

We found that data-driven decomposition of spectral power and connectivity of normal EEGs yields patterns that are interpretable in terms of known scalp electrophysiology and sensitive to physiological and pathological changes. Furthermore, the quantification of normal interictal EEG activity using these patterns revealed relevant group differences in focal epilepsy. These results suggest that quantitative characterization of normal interictal EEGs of focal epilepsy patients has the potential to augment visual EEG review and assist clinical decision-making in epilepsy. Future efforts will focus on validating these findings using a larger out-of-sample epilepsy cohort with data collected from an external site.

## 2. Data & Methods

### Clinical population dataset and expert EEG review

Our study utilized 13,652 routine clinical EEG recordings obtained from 12,134 adult patients (18 or older) at Mayo Clinic, Rochester, MN, USA between 2016 and 2022^25^. This study was approved by the Mayo Clinic institutional review board and patients provided informed consent. The EEGs were recorded using the XLTEK EMU40EX headbox manufactured by Natus Medical Incorporated, Oakville, Ontario, Canada. All EEGs followed the standard 10–20 electrode placement system^26^ and were sampled at 256Hz. The patient population comprises individuals presenting with a diverse array of conditions including epilepsy, cognitive impairment, episodic migraines, syncope, and functional spells, among others. Overall, this dataset represents the patient population typically referred for routine EEG assessments at the Mayo Clinic in Rochester, MN, USA. All EEG records were visually reviewed by board-certified epileptologists and graded based on the Mayo Clinic internal EEG grading protocol. EEGs within normal limits and without visible abnormalities were graded as normal. EEGs with asymmetry, persistent delta frequency slowing, and intermittent abnormalities were classified either as Dysrhythmia 1 (mild, non-specific slowing or excess of fast activity), Dysrhythmia 2 (moderate to severe intermittent slowing), or Dysrhythmia 3 (e.g. epileptiform abnormalities, triphasic waves, intermittent rhythmic delta frequency activity). Normal EEGs comprise the population set used for tensor decompositions. Note that patients corresponding to these normal EEGs may present with the aforementioned conditions including epilepsy.

### Focal epilepsy cohort and matched control subjects without epilepsy

Figure 1 depicts the process flow for constructing the epilepsy and control cohorts. Patients with EEGs containing focal epileptiform abnormalities (i.e., Dysrhythmia grade 3) were used to triage focal epilepsy cases in the overall patient population. Based on further review of those patients, we identified a total of 121 focal epilepsy patients (frontal=21; temporal=100; 125 EEGs) who had a confirmed diagnosis of frontal or temporal lobe epilepsy and had no prior history of any cranial surgery. The drug response status and MRI findings of patients with temporal lobe epilepsy were determined by reviewing electronic health records and diagnostic MRI reports available within a year of their EEG assessments, respectively. Cases where clinical evidence was either not available or insufficient were excluded from clinical sub-group classifications. Patients with frontal lobe epilepsy were not considered for these sub-group classifications due to low sample size. An age- and sex-matched control cohort of 76 subjects with normal EEGs and without diagnosis of epilepsy or other major neurological disorder was selected for comparisons from the overall set of normal EEGs. Data of patients in focal epilepsy and matched control sets were excluded from the population set during subsequent analyses to prevent statistical data leakage.

**Figure 1:**
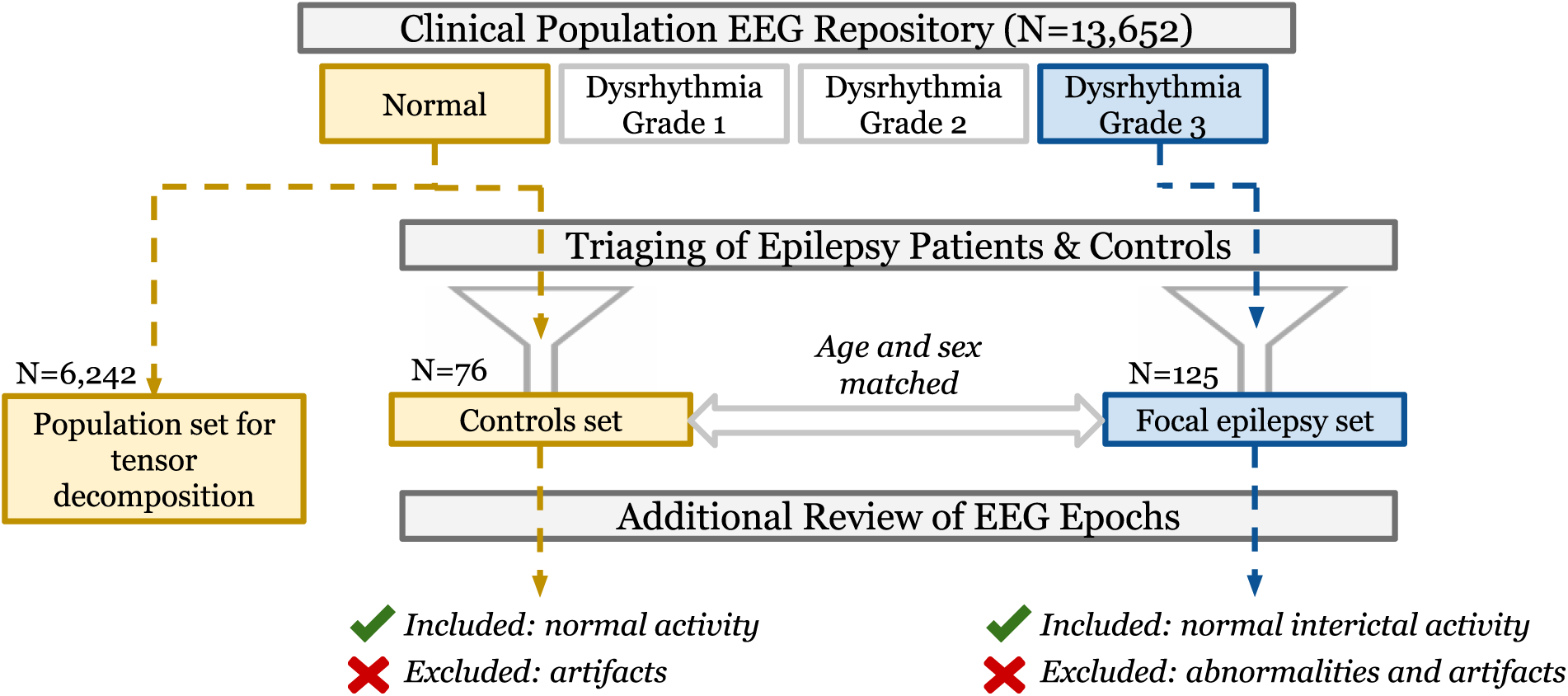
Cohort selection process flow starting from the overall clinical population dataset. Patients with focal epilepsy and controls without epilepsy were triaged using clinically assigned EEG grades, electronic health record notes/reports, and case reviews. Epochs extracted from their EEGs were reviewed for interictal abnormalities and excessive artifacts. Clinically graded normal EEGs comprise the population set for tensor decomposition.

The complete analytical workflow of this study from processing of raw EEGs to results is illustrated in Figure 2. Below we describe the methods used in this workflow.

**Figure 2:**
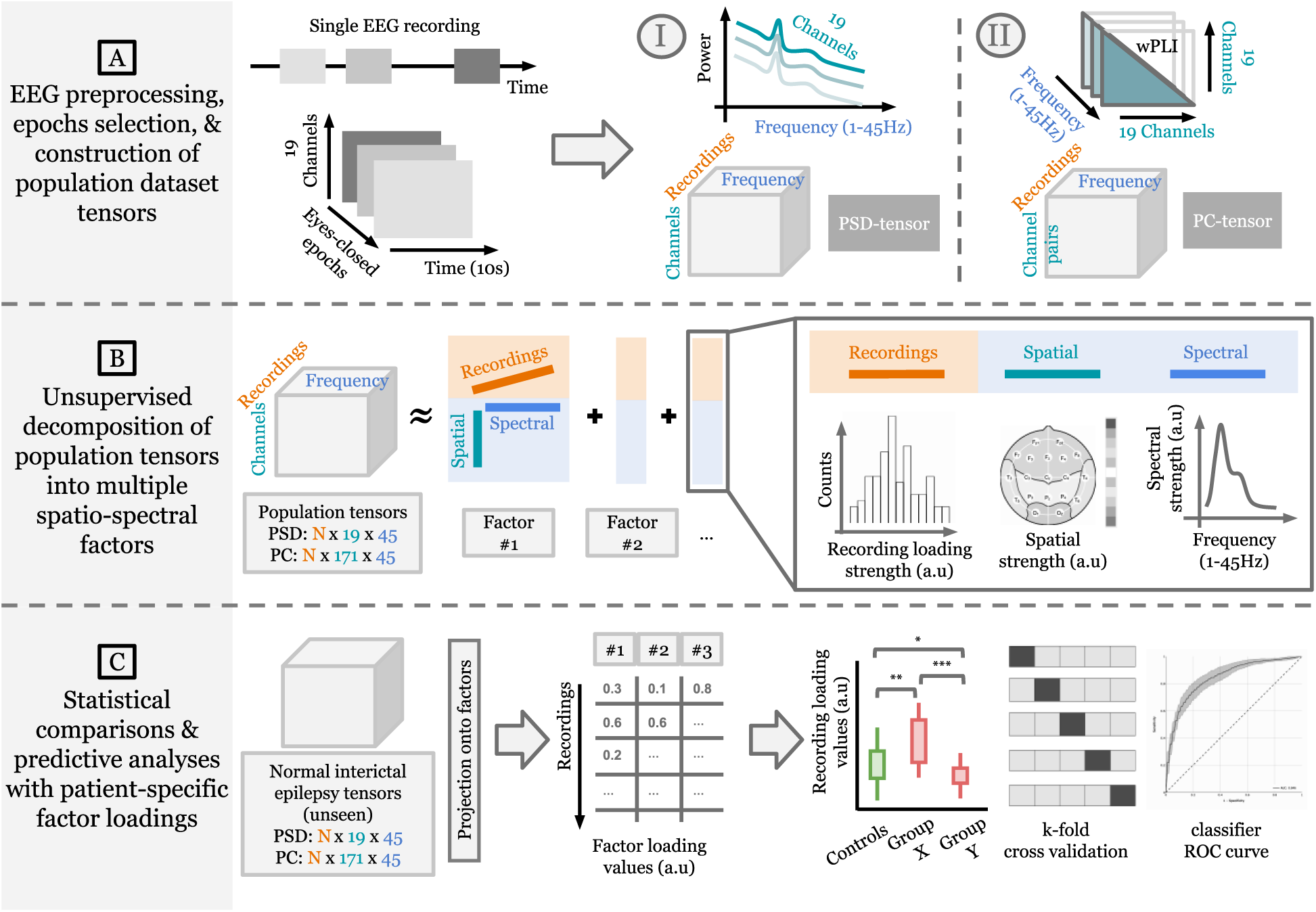
Overall analytic workflow of the study. (A) Multiple eyes-closed awake interictal epochs from each EEG recording are identified for data analysis. The average power spectral density (PSD) and phase-based connectivity (PC) between each channel pair are computed and stacked across recordings to obtain 3-d PSD and PC tensors (recordings x channels or channel pairs x frequencies). (B) PSD and PC population tensors are decomposed separately in an unsupervised fashion to obtain multiple interpretable spatio-spectral patterns (i.e., factors). (C) Normal interictal EEG data from focal epilepsy patients are projected on each population-level factor to obtain patient-specific factor loadings. Differences in drug-resistant and non-lesional MRI focal epilepsy are investigated by using these loadings in statistical group/sub-group comparisons and predictive analyses.

### EEG preprocessing and epochs selection

All routine EEGs were preprocessed as follows: 1) selection and ordering of the 19 EEG channels arranged according to the 10-20 system (i.e., Fp1, F3, F7, C3, T7, P3, P7, O1, Fp2, F4, F8, C4, T8, P4, P8, O2, Fz, Cz, and Pz), 2) resampling to ensure a sampling rate of 256 Hz, 3) band-pass filtering between 0.1-45Hz, and 4) transformation to common average reference. Artifact rejection was not performed in this pipeline as we hoped to recover population patterns specific to artifacts in a data-driven manner using tensor decompositions. Next, we applied a heuristic algorithm^27^ to select a maximum of six 10-second EEG epochs from the full recording representing eyes-closed wakefulness. The algorithm relies on sleep staging^28^, eye blinks, sample entropy, and occipital alpha power to select candidate epochs. These selected epochs are not guaranteed to be contiguous. After preprocessing, all EEG recordings were represented by at most six EEG epochs representing eyes-closed resting-state wakefulness. Preprocessing was done using the numpy^29^ and MNE^30^ Python libraries. Epochs selection used the MNE-features^31^ and YASA^32^ libraries.

### Additional review of EEG epochs extracted from focal epilepsy and control patients

From the extracted EEG epochs of focal epilepsy patients, a board-certified epileptologist visually reviewed and selected ones containing normal interictal activity. Abnormal epochs containing seizures, epileptiform spikes, epileptiform sharp waves, temporal intermittent rhythmic delta activity (TIRDA), and excessive artifacts were excluded from the study. Polymorphic, intermittent delta and theta frequency slowing (0.1 - <8 Hz) events, however, could not be excluded due to their pervasive presence in some EEGs. Similarly, epochs from non-epileptic controls with excessive artifacts were also excluded. We note that this additional review of epochs extracted using the automated algorithm was conducted only for epilepsy and control EEGs.

### Constructing tensors of spectral power

Power spectral density (PSD) of EEG data was estimated for all 19 EEG channels using Welch’s algorithm^33^, yielding log-power values at all integer frequencies between 1-45Hz. We then averaged the PSD measures of each EEG recording across all the identified epochs to obtain a single PSD vector for each channel. The PSD measures of each EEG recording can now be represented as a matrix with shape 19 × 45 (19 channels and 45 frequencies). Stacking this average PSD matrix across recordings produces a 3-d power-spectral tensor (“PSD-tensor”) of the form: N recordings x 19 channels x 45 frequencies. The population PSD-tensor is globally min-max scaled between [0, 1] to maintain non-negativity for subsequent tensor decomposition. Focal epilepsy and control PSD-tensors are scaled similarly but are stacked together first to preserve group differences for downstream analyses.

### Constructing tensors of phase-based connectivity

An estimate of phase-based connectivity (PC) between a pair of channels (*i*, *j*) is computed using the weighted Phase Lag Index^34^ (wPLI) measure defined as:

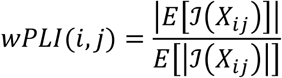

where *X*_*i*,*j*_ denotes the cross-spectral density of channels *i* and *j*, ℐ(. ) is the imaginary part of the cross-spectrum, and *E*[. ] represents a mean over the selected eyes-closed epochs. wPLI values range between [0, 1]. A positive value reflects an imbalance between leading and lagging relationships, with 1 indicating a perfect lead or lag relationship. At each integer frequency between 1-45Hz, wPLI provides a connectivity value for each of the 171 unique channel pairs. Thus, we obtain a 3-d phase-based connectivity tensor (“PC-tensor”) of the form: N recordings x 171 channel pairs x 45 frequencies.

### Representing the normal EEGs as 3-d population tensors

We utilized the clinically graded normal EEGs in the overall population dataset (N=6,242 out of 13,652) to extract population-level EEG patterns. We estimated the PSD and PC measures for these normal EEGs using their automatically extracted epochs and formed the population PSD-tensor and PC-tensor of shape (6,242 x 19 x 45) and (6,242 x 171 x 45), respectively.

### Decomposition of 3-d tensors into factors

The canonical polyadic (CP) decomposition^35,36^ (also known as the PARAFAC decomposition^37^) approximates a given tensor as a sum of *R* rank-1 tensors, where *R* is the decomposition rank, i.e., the resulting number of factors obtained from decomposing the tensor. The CP decomposition of a 3-dimensional tensor Τ with rank *R* is defined as:

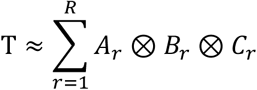

where ⊗ denotes an outer product and *A*_*r*_, *B*_*r*_, and *C*_*r*_ are vectors with shapes matching each of the three dimensions of Τ (recording, channel, frequency). Each term in the summation, i.e., a combination of *A*_*r*_, *B*_*r*_, and *C*_*r*_, is a rank-1 tensor and is referred to as a factor. The *A*, *B*, and *C* factor matrices (containing *A*_*r*_, *B*_*r*_, and *C*_*r*_ vectors as columns, respectively) are optimized with a non-negativity constraint using the hierarchical alternating least squares^37,38^ approach.

### Determining the initialization and rank for CP decomposition

We provided a physiologically meaningful initialization and rank derived from PSD characteristics of healthy subjects to initialize the decomposition of the PSD-tensor. For this, we fit a parametric model of the EEG PSD, named FOOOF^39^ (“fitting oscillations and one over f”), to the eyes-closed trials in the MPI Leipzig Mind-Brain-Body dataset^36^ (N=207, 8 trials per subject, 60s trial duration). The FOOOF model segments the observed morphology of an EEG PSD into superimposed aperiodic (*L*) and oscillatory components (*G*_*n*_):

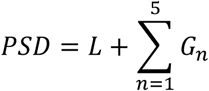

Each *G*_*n*_ is a Gaussian peak corresponds putatively to a canonical brain oscillation (delta, theta, alpha, beta, or gamma) and is parameterized by height, mean or center frequency, and a standard deviation. *L* is a function of the form 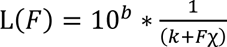 whose parameters *b*, *k*, and *χ* capture aperiodic 1/f-like nature of the *PSD*. We refer readers to Donoghue et. al. (2020) for additional model details. We fit this six-component model to healthy PSDs in the MPI-Leipzig dataset. The fitted versions of *G*_*n*_ and *L* formed the frequency initializations *B*_*r*_ of the decomposition solution and informed the choice of rank *R* = 6.

### Decomposing the population tensors

Factor matrix *B* (containing *B*_*r*_ vectors as columns) was initialized with the six spectral “priors” described above. CP decomposition with non-negativity constraints and *R*=6 was applied on the min-max scaled population PSD-tensor. The resultant *B* was then used as an immutable initialization for the subsequent CP decomposition of the population PC-tensor. In other words, only factor matrices *A* and *C* were optimized in the PC-tensor decomposition. The use of *B*, i.e., frequency patterns extracted from the PSD-tensor, in PC factors ensured that interpretations were aligned across both decompositions. Tensor analyses were done using the tensortools^40^ Python library.

### Visualization of factors derived from the normal EEG population

The *A*_*r*_, *B*_*r*_, and *C*_*r*_ vectors resulting from both CP decompositions represent semantically coherent components: *A*_*r*_ contains factor’s loadings per recording, *B*_*r*_ holds the factor’s channel activations, and *C*_*r*_ holds the factor’s frequency activations. The recording loadings are visualized as histograms, channel activations as topographical distributions over the scalp, and frequency activations as power spectral profiles. Note that we obtain *A*_*r*_ and *C*_*r*_ separately from the PSD-tensor and PC-tensor decompositions, while *B*_*r*_ is shared between both as described above. We refer to values in *A*_*r*_ as “PSD loadings” or “PC loadings” depending on the tensor they are associated with.

### Computing factor loadings for the focal epilepsy cohort

We computed population factor loadings for the focal epilepsy cohort using a projection operation^41^. Consider the basis matrix *P* containing vectorized versions of the spatio-spectral factors *B*_*r*_ ⊗ *C*_*r*_. Thus, matrix P has *R* rows and *C*\**F* columns, where *C* and *F* is the length of the channel dimension and frequency dimension of the tensor, respectively. Then, for a new EEG recording *x*_*new*_ ∈ *R*^*C*×*F*^, its loadings are computed by *P*^+^ × vectorized(*x*_*new*_), where *P*^+^ is the pseudo-inverse of *P*. The results of this operation are weights or loadings representing how strongly each factor is expressed in the new recording. Note that this operation does not guarantee non-negative loadings.

### Associations and statistical testing

Pearson’s correlation coefficient and Spearman’s rank correlation coefficient were used to quantify associations of factor loadings with patient age and ranked degree of slowing, respectively. The corresponding p-values test the null hypothesis that the distributions underlying the samples are uncorrelated. The Mann-Whitney-Wilcoxon two-sided test^24^ was used for group-level comparisons with Bonferroni correction^25^ for multiple comparisons. The test was performed using the stat-annot^26^ Python library.

### Predictive modeling

Patient-specific loadings were robustly scaled (subtract median, scale by interquartile range) and used as features in a logistic regression binary classifier. We explored three sets of features: PSD loadings, PC loadings, and both concatenated together. Nested k-fold cross-validation (CV) was done to assess variability of model performance on different held-out sets (outer CV loop, 10-fold) and to tune the ElasticNet regularization strength^45^ hyperparameter for each training set (inner CV loop, 5-fold). Grid for the hyperparameter search ranged between [0, 1] with increments of 0.1. Both CV loops used disjoint patient splits with target stratification. Loss values were weighted using target class proportions to handle class imbalance. For each outer CV fold, a classifier was trained using the best hyperparameter setting found by the inner CV loop and evaluated on the corresponding outer test fold. We used the area under receiver operating characteristic curve (AUC) to evaluate model performance across the outer CV folds. Predictive modeling was performed using the scikit-learn^46^ Python library.

### Data, code, and factor availability

Summary data and code can be made available by the corresponding authors upon reasonable request.

## 3. Results

### 3.1 Characteristics of the Neurological Population, Focal Epilepsy Cohort, and Controls

Table 1 provides an overview of the population-level routine EEG dataset. This dataset included 13,652 recordings from 12,134 unique patients. Expert visual review of these EEG recordings based on the Mayo Clinic grading criteria resulted in 45.7% (N=6,242) normal EEGs, 24.9% (N=3,395) EEGs with mild slowing (Dysrhythmia grade 1), 13.2% (N=1,800) EEGs with moderate to severe slowing (Dysrhythmia grade 2), and 16.2% (N=2,215) EEGs with epileptiform abnormalities (Dysrhythmia grade 3). From the population of Dysrhythmia grade 3 EEGs, we identified 121 focal epilepsy patients with clinically confirmed epilepsy in either the frontal (N=21) or temporal (N=100) region. In addition, a set of 76 matched non-epileptic controls with normal EEGs and without a diagnosis of any neurological disease were identified for group comparisons. Table 2 summarizes the characteristics of the confirmed epilepsy patients and controls.

**Table 1:** Characteristics of the overall neurologic clinical population.

| Data Property | Summary Statistics |
| --- | --- |
| Routine EEG recordings | Total recordings: 13,652<br>Unique patients: 12,134 |
| Age | Range: 18-103.7<br>Mean: 50.9 (± 19.4)<br>Age groups:<br>18 – 30: 2,639<br>30 – 50: 3,785<br>50 – 70: 4,563<br>>70: 2,665 |
| Sex | Female = 6,464 (53.3%) |
| EEG Grade (based on expert visual review) | Normal: 6,242 (45.7%)<br>Dysrhythmia 1: 3,395 (24.9%)<br>Dysrhythmia 2: 1,800 (13.2%)<br>Dysrhythmia 3: 2,215 (16.2%) |

**Table 2:** Characteristics of epilepsy cohort and controls used in this study.

| Study Cohort | Summary Statistics |
| --- | --- |
| Temporal Lobe Epilepsy (TLE) | Unique records: 100<br>Unique participants: 100<br>Age: 52.5 (19.9)<br>Sex: 50 (50%) Female<br>Drug response status:<br>44 Drug-resistant<br>28 Drug-responsive<br>28 Unknown<br>MRI status:<br>36 Non-lesional<br>43 Lesional<br>21 Unknown |
| Frontal Lobe Epilepsy (FLE) | Unique records: 25<br>Unique participants: 21<br>Age: 37.6 (13.6)<br>Sex: 12 (57.1%) Female |
| Non-epileptic Controls (CTL) | Unique records: 76<br>Unique participants: 76<br>Age: 49.2 (19.3)<br>Sex: 41 (53.9%) Female |

### 3.2 Tensor Decomposition Extracts Interpretable Spatio-spectral Patterns from Normal EEGs

Figure 3 shows the factors obtained by decomposing the normal EEGs in the population dataset, i.e., the population PSD-tensor and PC-tensor. The frequency profiles are largely distinct, except in the case of factors 2 and 6, where their spatial distributions uniquely characterize the overall pattern.

**Figure 3:**
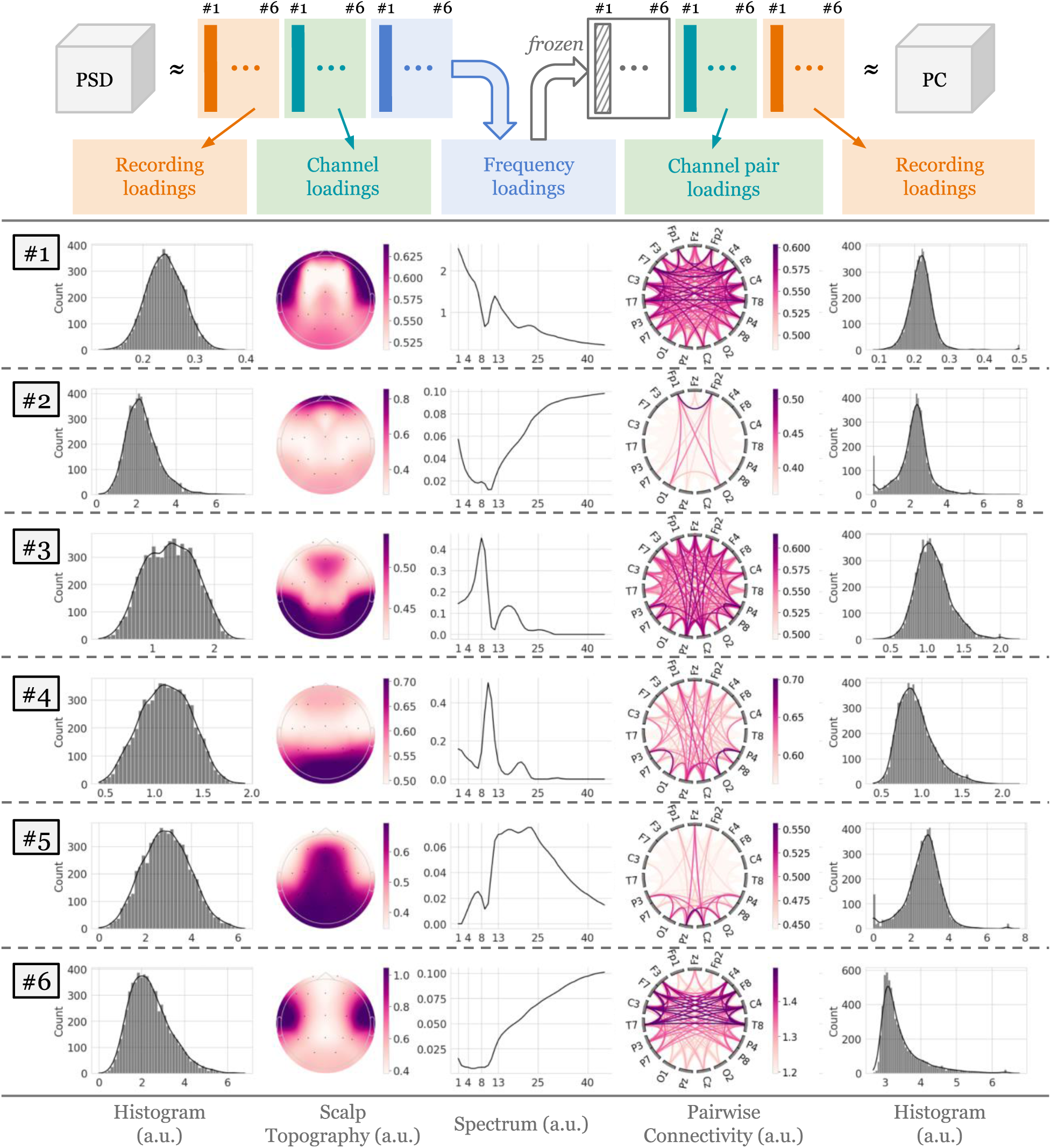
Data-driven population-level patterns of eyes-closed awake EEG data extracted from 6,242 normal EEGs. Three-dimensional tensors containing spatio-spectral information were decomposed using non-negative Canonical Polyadic Decomposition to yield six factors. Each row corresponds to a combination of a power spectral and connectivity-based factors, which is defined by the common spectral profile, the spatial power distribution over the 19 channels, the pair-wise channel connectivity, and loadings of EEG recordings in the PSD-tensor and PC-tensor. Recording loadings are visualized as histograms, spatial activations are visualized as scalp topographical distributions, and spectral activations are visualized as power spectral density. Note that the PSD-tensor was decomposed first, and the resulting frequency factors were kept frozen during the decomposition of the PC-tensor to align interpretation of the factors. (a.u. refers to absolute units.)

Factor 1 shows the characteristic 1/f frequency profile with minor deviations around the oscillatory bands and spatial activations in the fronto-temporal and posterior regions, characterizing the background non-oscillatory (i.e., aperiodic) brain activity. Factor 2 shows high frequency activations (>25Hz) in the prefrontal region, suggesting eye-movement-related artifacts. Factor 3 predominantly contains high-theta/low-alpha activity (6-9Hz) in fronto-parietal regions, possibly indicating the high theta rhythm or slow alpha rhythm. Factor 4 shows occipital activations in 8-13Hz, resembling the characteristic posterior dominant rhythm. Factor 5 shows centro-parietal activations in 13-25Hz, capturing the Rolandic beta activity. Lastly, factor 6 shows high-frequency activations (>25Hz) in the temporal regions, which may represent muscle artifacts. The analyses and findings presented in the remaining text focus on the four putatively physiologic factors (1, 3, 4, and 5).

### 3.3 Patient Loadings Show Sensitivity to Aging and EEG Dysrhythmia Grades

Figure 4 shows the associations between the loadings of population EEGs for factors 1, 3, 4, and 5 against patient age and expert-assigned EEG grades

**Figure 4:**
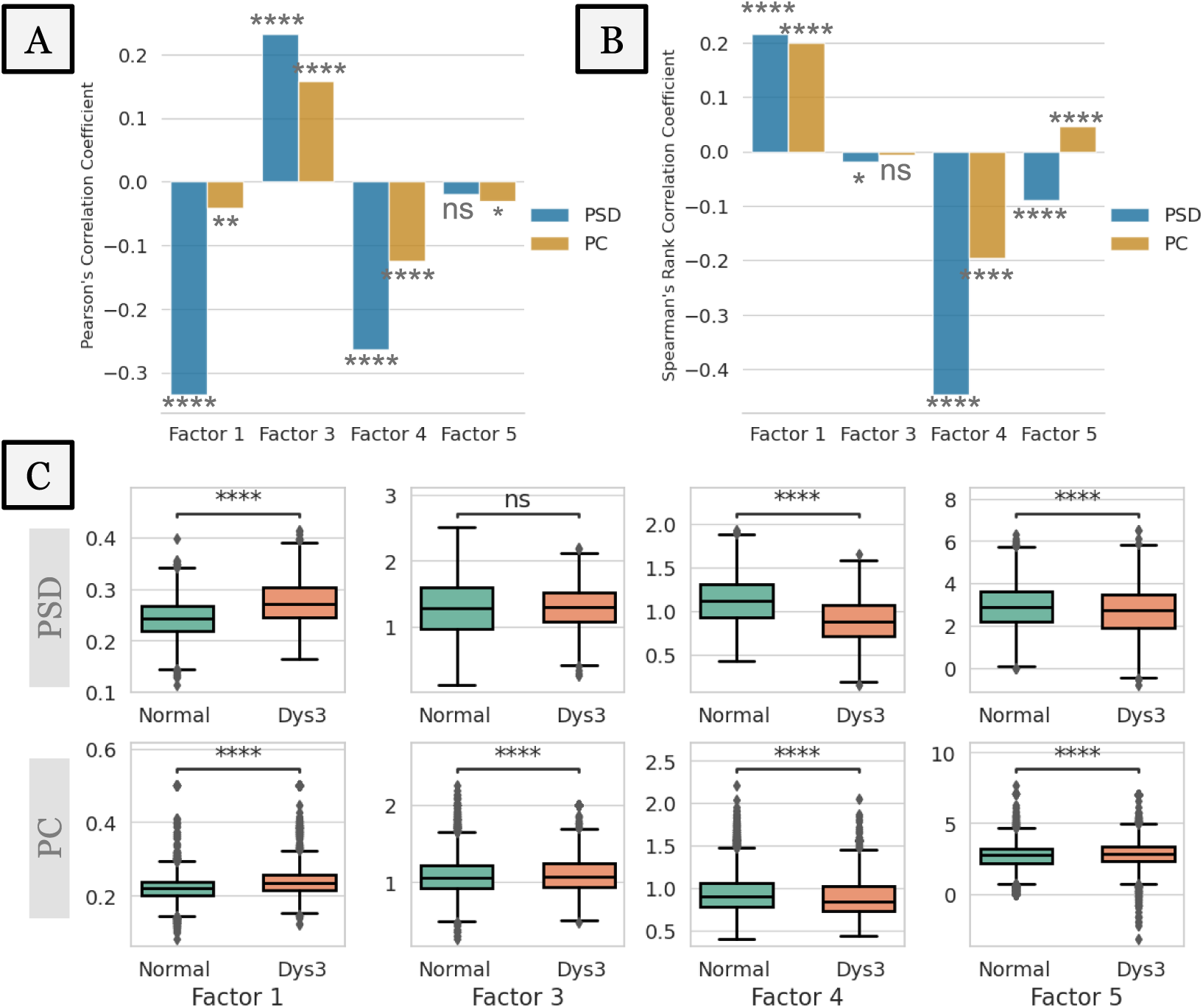
Associations of PSD and PC loadings of the four putatively physiologic factors (1, 3, 4, and 5) with physiological (aging) and pathological (slowing, epileptiform activity) variables. Factor numbers correspond to those in Figure 3. Loadings describe activity found in eyes-closed awake EEG segments selected from expertly graded routine EEGs in the population-level dataset. (A) Correlations of PSD and PC loadings of normal EEGs with patient age. (B) Correlations of PSD and PC recording loadings with expert-assigned severity of slowing. The ranked severity levels are 0 (normal EEG, no slowing), 1 (Dysrhythmia 1 EEG, mild slowing), and 2 (Dysrhythmia 2 EEG, moderate to severe slowing (C) Correlations of PSD and PC recording loadings with the presence of epileptiform activity (Dysrhythmia 3 EEGs abbreviated as “Dys3”). Significance levels correspond to the Mann-Whitney-Wilcoxon test. Loading values along y-axes are in arbitrary units. * indicates a significant correlation with p < 0.05 and **** indicates a significant correlation with p < 1e-4.

*Trends with patient age (*Fig. 4A): Factor 3 is positively correlated with age (p<1e-4), while factors 1 (PSD: p<1e-4, PC: p<0.01) and 4 (p<1e-4) are negatively correlated. Although the correlation strength varies between the PSD and PC loadings of the same factor, they are directionally consistent. Correlations of factor 5 are either marginally significant (PSD: p<0.05) or not significant (PC).

*Trends with expert-ranked degree of slowing (*Fig. 4B): Factor 1 is positively correlated with severity of slowing (p<1e-4), while factor 4 is negatively correlated (p<1e-4). Correlation of factor 3 is either low (PSD: p<0.05) or not significant (PC). The correlation of factor 5, although significant (p<1e-4), is directionally divergent between the PSD and PC loadings.

*Differences in presence of epileptiform activity (*Fig. 4C): Here, loadings of EEGs with epileptiform activity were compared against those of normal EEGs. PSD loadings of factor 1 increase under presence of epileptiform activity, while those of factors 4 and 5 decrease (p<1e-4 in every case). Factor 3 PSD loadings show no significant change. PC loadings of factors 1 and 4 show trends consistent with corresponding PSD loadings (p<1e-4 in both cases). However, the PC loadings of factors 3 and 5 show slight increases (p<1e-4).

### 3.4 Quantitative Analysis of Normal Interictal EEG Reveals Differences in Focal Epilepsy

Figure 5 shows results for group differences and binary classifications between non-epileptic controls and the focal epilepsy cohort using patient-specific PSD and PC loadings of the physiologic factors. We find focal epilepsy patients to have elevated factor 1 (p<0.001) and factor 3 (p<0.05). in both PSD and PC comparisons (Figure 5A-B). In addition, we find PC loadings for factor 5 (p<0.05) significantly different in focal epilepsy relative to non-epileptic controls. Factor 4 loadings do not show significant differences in either the PSD or PC comparisons.

**Figure 5:**
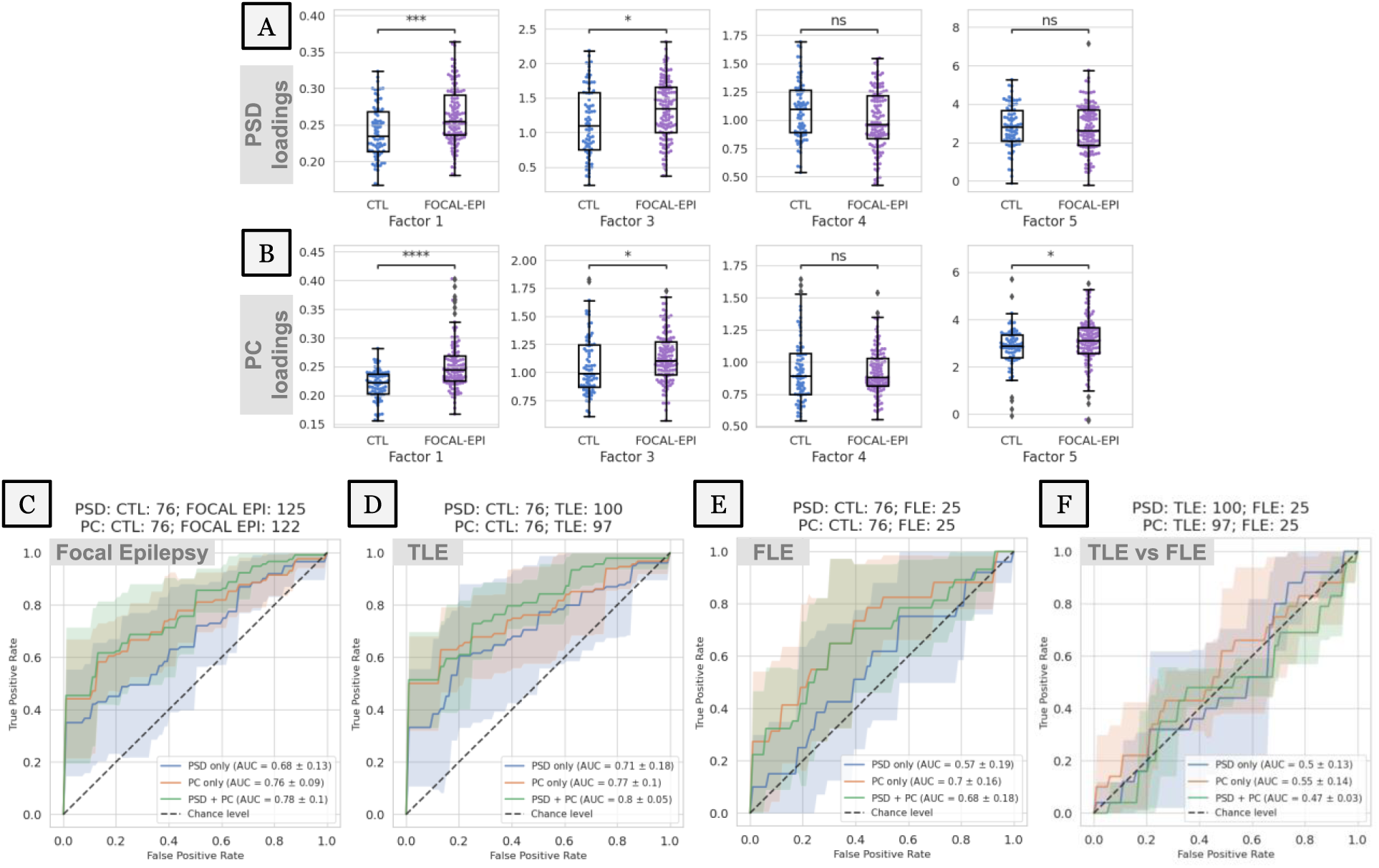
Differentiation of focal epilepsy and epileptogenic. (A-B) PSD and PC loadings of focal epilepsy patients (FOCAL-EPI) are compared to those of non-epileptic controls (CTL) across the four physiologic population factors. Loading values along y-axes are in arbitrary units. * indicates a significant difference with p < 0.05 and **** indicates a significant difference with p < 1e-4 in the Mann-Whitney-Wilcoxon test. (C) PSD and PC loadings are used as features to classify focal epilepsy vs. non-epileptic controls within a binary classification framework. (D-E) The same classification is broken down by temporal (TLE) and frontal (FLE) sub-types of focal epilepsy. (F) Differential diagnosis of the epileptogenic lobe, i.e., TLE vs. FLE, within the focal epilepsy cohort. Note that all classifications used only the four putative physiologic factors (1, 3, 4, and 5) and were conducted with three sets of features/loadings - only those of PSD factors (“PSD only”), only those of PC factors (“PC only”), or both concatenated (“PSD + PC”).

Figure 5C shows classification of focal epilepsy vs. non-epileptic patients is possible above chance levels, with PC loadings providing the largest contribution to the average classification performance (AUC=0.76). This performance is marginally improved by using a combination of PSD and PC loadings (AUC=0.78). All feature sets show high variability in performance across the held-out folds (0.09-0.13). Figures 5D-E show results for the classification of frontal (FLE) and temporal lobe epilepsy (TLE) against non-epileptic controls. TLE is better differentiated from non-epileptic patients than FLE (top mean AUC=0.8 vs. 0.7). TLE is best differentiated by combined PSD and PC loadings (AUC=0.80), with PC loadings contributing the most to classifier performance (AUC=0.77). FLE is best differentiated using PC loadings alone (AUC=0.70), and the addition of PSD loadings slightly worsens the performance (AUC=0.68). Variability in AUC performance across folds ranges from 0.05-0.19. Lastly, Figure 5F shows the classification of TLE vs FLE based on factor loadings derived from normal interictal epochs. Results indicate that none of the feature sets can differentiate the epileptogenic lobe (i.e., temporal vs. frontal) in focal epilepsy above chance levels (AUCs range between 0.47-0.55) based on normal interictal epochs.

### 3.5 Quantitative Loadings of Normal Interictal EEG Exhibit Capacity for Differentiation in Drug-Resistant and Non-lesional Epilepsy

Figure 6A shows differences in loadings of non-epileptic controls (CTL), drug-responsive (TLE-respon), and drug-resistant (TLE-resis) temporal epilepsy patients. Only the PSD loadings for factor 5 show differences between the two sub-groups (p<0.05), while the others show differences only relative to controls. None of the PC loadings show significant differences between the two sub-groups. PC loadings other than those of factor 1 show no differences between non-epileptic controls and both sub-groups. Figure 6B shows the classification performance of different sets of factor loadings in classifying drug resistance. PSD loadings provided the best average performance (AUC=0.73) while PC loadings performed marginally better than chance (AUC=0.58). Variability in model performance ranged from 0.07 to 0.13 AUC points.

**Figure 6:**
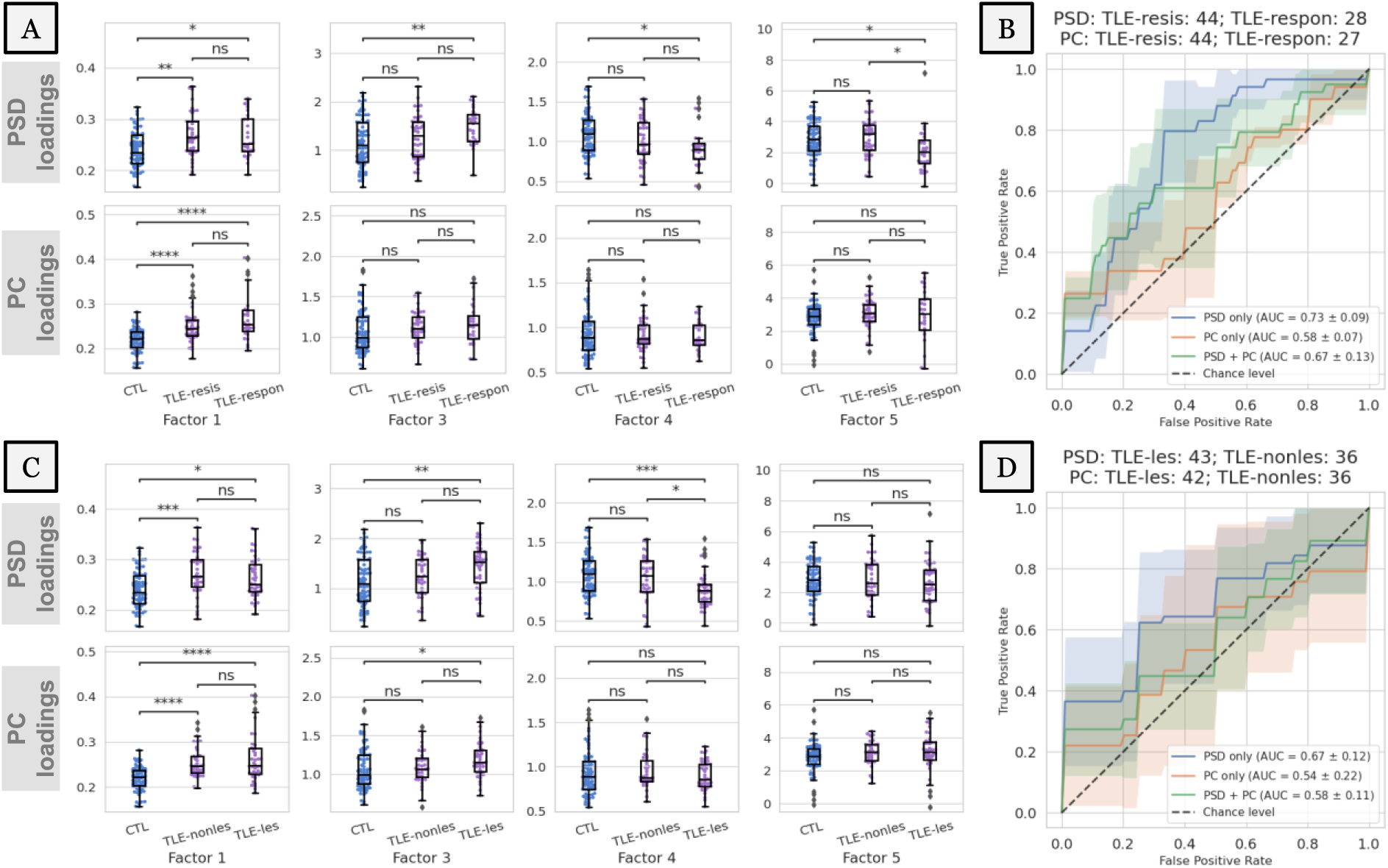
Differentiation of drug-resistant and non-lesional temporal lobe epilepsy (TLE) patients using four physiologic pattern loadings (factors 1, 3, 4, and 5). (A) Loadings are compared between non-epileptic controls (CTL), TLE patients that are drug resistant (TLE-resis) and those that are drug responsive (TLE-respon). (B) Binary classifications of drug resistant vs. responsive patients using the same feature sets as Figure 5. (C-D) Analyses similar to (A) and (B) are conducted for lesional (TLE-les) and non-lesional (TLE-nonles) TLE sub-groups. Loading values in (A) and (C) along y-axes are in arbitrary units. * indicates a significant difference with p < 0.05 and **** indicates a significant difference with p < 1e-4 in the Mann-Whitney-Wilcoxon test with Bonferroni correction.

Figure 6C shows differences in normal interictal EEG loadings between non-epileptic controls (CTL), non-lesional (TLE-nonles), and lesional (TLE-les) temporal lobe epilepsy. While PSD loadings of factors 1, 3, and 4 show significant differences relative to non-epileptic controls for both groups, only factor 4 shows a significant difference between non-lesional and lesional patients (p<0.05). Trends seen in factors 1 and 3 are similar between the PSD and PC loadings. However, none of the PC loadings differed significantly between the MRI sub-groups. Figure 6D shows the classification between lesional and non-lesional patients. PSD loadings best differentiate the two groups of patients with an AUC of 0.67. PC loadings, either alone or in addition to PSD loadings, significantly worsened the average classification performance. However, all models exhibited high variability in AUC performance (0.11-0.22 AUC points).

## 4. Discussion

The goal of this study was to explore whether normal interictal EEGs of people with focal epilepsy contain subtle signals that could be used to augment epilepsy diagnosis and treatment planning, especially in patients with drug-resistant and MRI normal epilepsy. We proposed a scalable, physiology-informed, and data-driven tensor decomposition approach that extracts spatio-spectral patterns from a large population of normal routine EEGs. Each pattern had a distinct signature in the EEG channel (spatial) and frequency (spectral) dimensions. We obtained patient-specific pattern loadings or “features” that allowed us to study group differences through statistical comparisons and binary classifications. Our findings suggest that quantitative description and analysis of visually reviewed normal routine EEGs has the potential to provide additional value to clinical decision-making in epilepsy.

### Tensor Decomposition with Spectral Priors Recovers Interpretable Patterns

This study hypothesized that the information content of normal EEGs can be explained by a parsimonious number of latent patterns. To test this hypothesis, we decomposed the spectral and connectivity contents of a population of normal routine EEGs into several meaningful patterns (i.e., factors) using a canonical polyadic tensor decomposition. In general, determining the exact number of factors, i.e., the presumed rank of the population tensor, is challenging and involves trial-and-error^47^. However, prior work has demonstrated that the morphological content of the scalp EEG PSD can be sufficiently explained by six physiological components, namely one aperiodic 1/f pattern and five oscillatory bands^39^. We used this spectral parameterization model to construct six corresponding frequency priors that, in turn, provided the spectral initialization as well as an appropriate rank for the decomposition. Furthermore, we fixed the spectral patterns extracted from PSD-tensor during the decomposition of PC-tensor to recover semantically consistent patterns from both the tensor types.

Several prior works have explored data-driven or unsupervised recovery of spatial, spectral, or temporal profiles of oscillatory sources and background patterns comprising spontaneous EEG activity^48–52^. In this study, we presented an approach that quantifies spatio-spectral EEG patterns with the goal of decision support when clinical EEGs are normal on expert visual review. Beyond the use of spectral-prior-based initialization, our approach did not place any assumptions on the statistical nature or morphology of the latent EEG patterns and can be applied without sophisticated artifact removal.

The population patterns (Fig. 3) can be loosely interpreted to reflect dominant and overlapping physiological processes whose linear superposition (summation) yields the original EEG trace. We then interpreted the identified patterns based on clinical domain knowledge. The putative interpretations of these patterns are supported by their sensitivity to patient age and severity of pathology (Fig. 4).

### Augmenting Epilepsy Diagnosis and Treatment Planning

Scalp EEG is an indispensable tool in epilepsy that can non-invasively record brain electrical activity with excellent temporal resolution. Due to this unique resolution, scalp EEG tests can capture transient interictal epileptiform discharges (IEDs) such as epileptiform spikes or sharp waves associated with epilepsy^53^. In current clinical practice, the expert identification and characterization of IEDs on routine scalp EEG is crucial for epilepsy diagnosis. Routine EEGs are also useful in measuring the efficacy of ongoing ASM trials^54^. In the case of drug-resistant epilepsy, the distribution of IEDs identified on scalp EEGs can help localize the seizure onset zone, especially in patients with no visible lesion on MRI. Thus, the identification of IEDs is central to the clinical value of scalp EEGs in current practice.

Recent studies have shown significant interest in the automated identification of IEDs to augment expert visual review ^24,55,56^. However, the diagnostic yield of a single routine scalp EEG is limited, with only 29-55% of them capturing epileptiform abnormalities^57^. Multiple EEGs may increase epileptiform yield up to ∼75%^58,59^, but the expected gain sharply drops after the third normal EEG. As such, normal interictal EEGs can cause treatment delays in multiple stages of epilepsy care. Previous studies that explored biomarkers of interictal non-epileptiform EEG support the possibility of augmenting decision support in epilepsy using spectral and connectivity-based EEG features^17–19,27,60–64^. Drawing inspiration from these smaller scale studies, we explored data-driven recovery of spectral features using a large population dataset of normal EEGs and analyzed their differences in epilepsy.

Our findings in Figures 5 and 6 suggest that normal interictal EEG activity of focal epilepsy patients contains significant differences in putative physiologic oscillations (factors 3, 4, and 5) as well as aperiodic 1/f(Hz) activity (factor 1). Increases in expression of 1/f and theta frequency activity, coupled with a decrease in alpha frequency may represent general intermittent slowing of the EEG background. Although we identified differences in factor 5, the differences in beta frequency rhythm may arise due to the presence of ASMs. The factors exhibited relatively lower performance in detecting FLE (Fig. 5E) and in differentiating FLE vs TLE (Fig. 5F). We believe that this may be due to either the lower sample size of the FLE cohort compared to the TLE cohort (Fig. 5D) or the global/symmetric nature of the population patterns.

### Understanding Subtle Variation in Visibly Normal EEGs through their Quantitative Descriptors

Our results (Figure 5) indicate that factor loadings extracted from normal EEG segments have the potential to classify focal epilepsy above chance levels (best mean AUC=0.78). We analyzed the changes in actual power spectral and timeseries data corresponding to the changes in factor loadings to further illuminate the factor interpretations.

Fig. 7A shows the full power spectra of normal EEG segments whose loadings fall in the bottom 10-percentile (low), between 40-60-percentile (medium) and top 10-percentile (high) of a particular physiologic oscillatory factor. We find that EEGs that score high in factors 3, 4, and 5 have higher power in high-theta/low-alpha, alpha, and beta bands, respectively.

**Figure 7:**
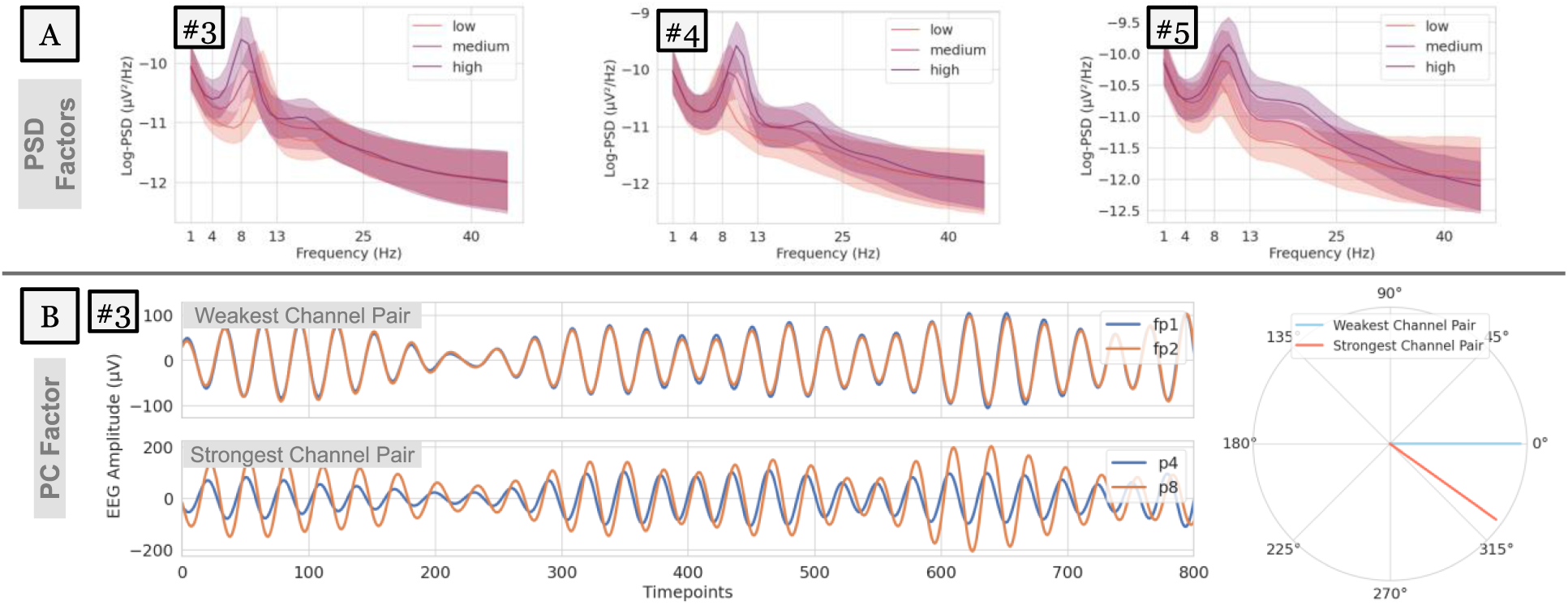
Variability in EEG power and phase characteristics based on factor loading values. (A) Variability in the power spectra of EEGs whose PSD loadings score in the bottom 10-percentile (low), between 40-60-percentile (medium), and top 10-percentile (high). Examples are shown for factors 3, 4, and 5. (B) 8-Hz-filtered EEG traces of the weakest (top) and strongest (bottom) channel pairs for an example EEG that scored in the top 10-percentile for factor 3 (whose spectral power peaks at 8Hz). Overlapping EEG traces reveal phase relationships, i.e., time lags that maximize correlation within the channel pairs. These lags or phase differences are visualized in polar coordinates (right).

Effects of the phase-lag-based connectivity (i.e., wPLI) at a particular frequency can be observed by leading/lagging relationships in the time-domain EEG signal filtered at that frequency. Fig. 7B focuses on factor 3 whose spectral power peaks at 8Hz, with the weakest edge connecting Fp1 and Fp2, and the strongest edge connecting P4 and P8 (shown in Figure 3). We visualize the phase relationships using an example EEG segment whose loading value was in the top 10-percentile for factor 3 after filtering its EEG trace around 8-Hz to. We find that the strongest channel pair (Fig 7B, bottom) has a consistent non-zero phase difference, while the weakest channel pair (Fig 7B, top) has no phase difference. These phase differences can be quantified by the time lag that maximizes timeseries correlation within the channel pair and are visualized in polar coordinates (Fig 7B, right).

These illustrations highlight that the quantitative loading values provided by this tensor-based framework are interpretable based on physiologically relevant concepts such as signal power and phase and offer sensitivity to subtle changes in the EEG signal. These subtle changes in normal EEGs are likely to be missed during traditional expert visual review, which focuses mostly on transient abnormalities in the time domain.

### Influence of Sample Size and Selected EEG Epochs on Study Findings

The routine EEG protocol contained diverse patient states (eyes-closed, eyes-open, awake, drowsy, asleep) and provocative maneuvers^65^ (photic stimulation, hyperventilation, sleep deprivation), making it necessary to select EEG epochs corresponding to a fixed patient state for data analysis. Such data selection may introduce bias in our findings since we selected only a maximum of six EEG epochs from each recording for our analyses.

To evaluate whether a bias exists, we repeated the controls vs TLE classification (result in Fig. 5D) with two bootstrapping strategies, whose results are shown in Figure 8. In strategy A (Fig. 8A), we considered TLE patients (N=41) with exactly six normal interictal EEG epochs and showed differences in classification performance depending on which 50% data are used for classification (i.e., first three epochs or last three epochs). Mean performance was higher when the first 3 epochs were used (AUC=0.65) than last 3 epochs (AUC=0.59). In strategy B (Fig. 8B), we maintained the sample size of the original TLE cohort (N=100) but used at most three randomly picked EEG epochs per recording to perform classification. For patients with >3 epochs available, 3 epochs were randomly chosen and for those patients with <=3 epochs, all epochs were chosen. Our results did not show any significant differences between those two sampling approaches and the overall performance closely matched that using all available epochs.

**Figure 8:**
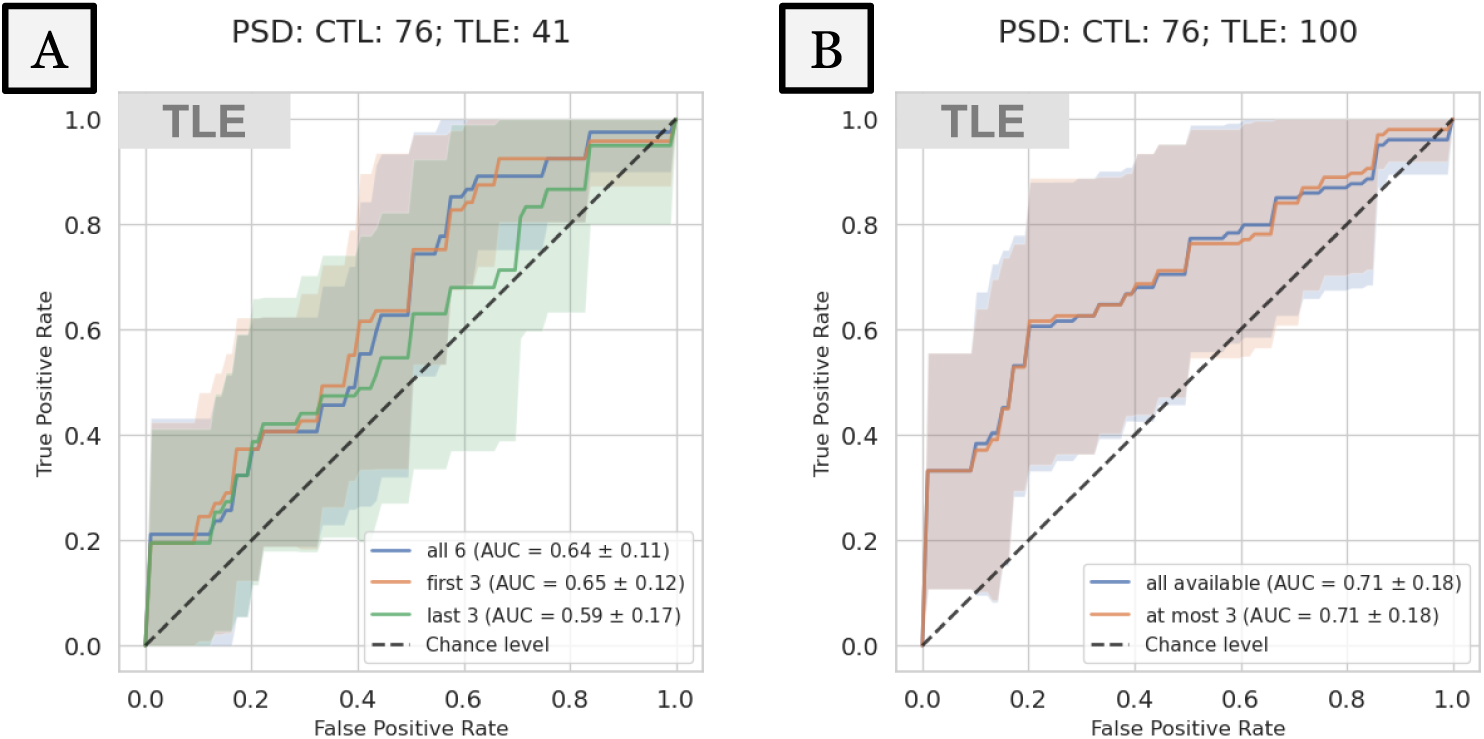
Repeated CTL vs TLE classifications using two bootstraps to evaluate bias introduced by the dataset selection process. Strategy A (left) uses either the first or last three of the six EEG epochs from a subset of TLE patients (N=41). Strategy B (right) uses at most 3 epochs that are randomly chosen but uses all available TLE patients (N=100).

These results suggest that: 1) our findings may be sensitive to low cohort size but are less likely to be biased by the algorithmic selection of EEG epochs within a recording, and 2) even as few as three normal interictal EEG epochs (30 seconds) are sufficient to derive a pretest measure of TLE.

### Study Limitations

Our goal in this study was to evaluate whether a quantitative analysis of normal EEG segments of epilepsy patients can indicate the possible presence of focal epilepsy. To test this hypothesis, we analyzed non-epileptiform interictal segments identified by a board-certified epileptologist within EEG recordings containing epileptiform abnormalities at other times (i.e., Dysrhythmia grade 3). However, an analysis using entirely normal EEGs of epilepsy patients will be necessary to evaluate the true potential of our results. However, identification of such EEGs requires extensive review of patient records, which we hope to accomplish in a follow-up study. Furthermore, eyes-closed wakefulness was determined by a heuristic algorithm validated in previous studies^27,66^. Events markers or comments added by EEG technologists^70^ during the EEG study could help to identify the patient’s behavioral state more reliably. Extension of our analysis to different sleep states will be pursued in future studies.

The estimation of connectivity could benefit from EEG source modeling to avoid volume conduction^71^ and active reference^72^ effects on the scalp. However, the lower spatial density of clinical EEGs prevented source/inverse modeling efforts, as previous studies have shown that EEG source modeling with fewer than 64 channels is highly error-prone^68–70^. Phase-based connectivity, and wPLI in particular, was chosen to suppress spurious zero-lag correlations and partially alleviate the effects of volume conduction^67,68^. Due to absence of patient-specific head models, average referencing was chosen to mitigate reference-related effects on connectivity better than alternatives like Cz and linked mastoids^69^.

Our classification analyses demonstrated a high level of variance between cross-validation folds (Fig. 5 and Fig. 6). Such variance could be a result of low sample size and the potential effects of comorbidities^70,71^ and medications^72^. The effects of these confounders may be mitigated either by comprehensive patient review to identify a clinically homogeneous set of focal epilepsy patients or with the use of larger epilepsy and matched control cohorts. Given that the EEG background patterns identified in this study are not specific to epilepsy, apparent differences in factor loadings must be interpreted within the appropriate clinical context. Additionally, validations using normal interictal EEGs from an external site are needed to assess the generalizability of the presented findings.

## 5. Conclusion

Normal interictal EEGs recorded from epilepsy patients can lead to delays in neurological care, especially in patients with drug-resistant and normal MRI epilepsy. This study explored the value of quantitative analysis of normal interictal EEGs in supporting a focal epilepsy diagnosis. Application of this unsupervised learning approach could benefit treatment planning in the future. We presented a scalable, interpretable, data-driven approach based on canonical polyadic decomposition that recovered physiologically meaningful spectral power and phase-based connectivity patterns from a population-scale dataset of normal EEGs and provided patient-specific loadings for each pattern. These loadings demonstrated value in classifying focal epilepsy and, in temporal lobe epilepsy, drug resistance and absence of lesions. These findings suggest that normal routine EEGs may contain subtle abnormalities that can be captured using a quantitative approach and be potentially used to augment decision-making in clinically challenging scenarios.

## Data Availability

Summary data and code can be made available by the corresponding authors upon reasonable request.

## Acknowledgements

We thank the Mayo Clinic Neurology Artificial Intelligence Program (NAIP) for guidance on the data processing workflow.

